# Consistent Differential Effects of Bupropion and Mirtazapine in Major Depression

**DOI:** 10.1101/2024.12.27.24319612

**Authors:** Eric V. Strobl

## Abstract

**Background:** Patients with major depression exhibit a wide range of responses to antidepressants. Unfortunately, most clinical trials fail to differentiate the effects of treatments on the primary symptoms of major depression, partially because they rely on fixed outcome measures such as total symptom severity scores or remission rates.

**Methods:** We performed a comprehensive analysis of the STAR*D trial with the Supervised Varimax (SV) algorithm incorporating post-model selection inference in order to *learn* outcome measures that differentiate between antidepressants. We also ran the algorithm on an independent clinical trial called CO-MED.

**Outcomes:** We differentiated bupropion and mirtazapine from multiple other antidepressants in STAR*D with replication in every relevant trial level. We further differentiated bupropion augmentation from mirtazapine augmentation in CO-MED. In particular, bupropion monotherapy had a greater therapeutic effect on hypersomnia than venlafaxine monotherapy in Levels 2 and 2A of STAR*D (*n* = 686, difference = 0.384, *p*_*FWER*_ = 0.007). Bupropion augmentation outperformed buspirone augmentation in Level 2, especially in patients with increased weight, increased appetite and fatigue (*n* = 520, difference = −0.322, *p*_*FWER*_ = 0.005). In contrast, mirtazapine monotherapy had greater therapeutic effects on insomnia, decreased weight and decreased appetite than nortriptyline monotherapy in Level 3 (*n* = 214, difference = 0.401, *p*_*FWER*_ = 0.022). Similarly, venlafaxine with mirtazapine augmentation out-performed tranylcypromine in Level 4, especially in patients with insomnia, decreased weight and decreased appetite (*n* = 102, difference = −0.722, *p*_*FWER*_ = 0.004). Finally, escitalopram with bupropion had larger therapeutic effects on increased weight, increased appetite and fatigue in CO-MED, while venlafaxine with mirtazapine had larger therapeutic effects on decreased weight, decreased appetite and insomnia (*n* = 640, difference = −0.302, *p*_*FWER*_ = 0.022).

**Interpretation:** Bupropion monotherapy and augmentation are effective specifically when a patient suffers from hypersomnia, increased weight, increased appetite or fatigue. Mirtazapine monotherapy and augmentation are effective in the opposite scenario, when a patient suffers from insomnia, decreased weight or decreased appetite.

## Introduction

Many clinical trials in psychiatry struggle to differentiate treatment from placebo, let alone treatment from a second active treatment. As a result, only a handful of clinical trials have identified differential effects between *active* psychiatric treatments. The problem is particularly pronounced in major depression, where nearly every clinical trial has failed to identify significant differences between antidepressants, despite some trials recruiting hundreds of patients. For example, Level 2 of the STAR*D trial involved over 1400 patients, but all antidepressants yielded near identical average response curves [1]. Consistently differentiating antidepressants in clinical trials therefore remains a grand challenge in psychiatry.

Investigators have thus far attempted to differentiate antidepressants (and other treatments) by adopting one of two general strategies. The first approach increases the sample size to tens, if not hundreds, of thousands of patients by combining many clinical trials in meta-analyses [2, 3, 4, 5]. These studies ultimately differentiate treatments at statistically significant thresholds due to the sheer number of patients, but with only marginally different effect sizes of limited clinical utility. Waiting for the results of multiple clinical trials can also slow down the advance in precision psychiatry. The second approach trains complex predictive models that utilize many baseline biological or clinical characteristics to predict depression severity or remission status (e.g., [6, 7, 8, 9]). These methods thus attempt to differentiate treatment response by fine-grained patient stratification. Unfortunately, increased complexity also increases fragility, where even slight changes in the patient population, such as in different hospitals or in different clinical trials, can dramatically decrease performance to chance level [10]. Deploying, generalizing and maintaining predictive models therefore requires substantial human effort in practice.

Psychiatrists do not differentiate medications by attempting to decrease total severity scores in specific subpopulations like predictive models. Instead, psychiatrists often differentiate treatments by memorizing their unique *effects* on individual symptoms, i.e., the individual items in an outcome rating scale. For example, a psychiatrist may prescribe mirtazapine to a patient with major depression experiencing insomnia and weight loss, because mirtazapine promotes sleep and increases appetite. Notice that the psychiatrist chooses mirtazapine because of its unique effects on sleep and appetite, rather than because of its predicted effect on a total depression severity score or remission status in a patient subgroup. We, therefore, recently proposed the Supervised Varimax (SV) algorithm that mimics psychiatrists by learning the outcome measures that best differentiate between treatments [11]. SV takes the individual items in a clinical rating scale as input and outputs sub-scales involving only the primary symptoms that specific treatments target well, such as insomnia and weight loss in major depression for mirtazapine.

In this paper, we perform a comprehensive analysis of the STAR*D trial using the SV algorithm in order to identify optimal outcome measures that differentiate antidepressants. We hypothesized that the algorithm would differentiate bupropion, or the most activating antidepressant tested, from mirtazapine, or the most sedating antidepressant tested. Furthermore, SV would replicate the differential effect in all sub-trials (or levels) within STAR*D, and in a completely independent clinical trial called CO-MED.

## Materials and Methods

### Clinical Trials

We analyzed two clinical trials of major depression. The trials have been described in detail elsewhere. We describe the parts of the clinical trials relevant to this manuscript below.

1. Sequenced Treatment Alternatives to Relieve Depression (STAR*D, ClinicalTrials.gov, NCT00021528, [12, 13]) was a double-blind, randomized and multi-center clinical trial that compared the efficacy of different sequences of antidepressants in major depressive disorder. The trial had multiple levels listed below: Level 1: All patients received citalopram. Level 2: Patients progressed to Level 2 if they failed to achieve remission in Level 1 and agreed to at least one of the following four options: medication switch, medication augmentation, cognitive therapy switch, and cognitive therapy augmentation. Patients then underwent randomization among the treatment options that they agreed to. As a result, patients only underwent strict randomization among (a) the medication switch options including bupropion, sertraline and venlafaxine, as well as (b) the medication augmentation options including buspirone augmentation and bupropion augmentation. Level 2A: Patients progressed to Level 2A if they received cognitive therapy switch or cognitive therapy augmentation in Level 2 but failed to achieve remission. Patients were randomized to either bupropion or venlafaxine monotherapy. Level 3: Patients progressed to Level 3 if they failed to achieve remission in Level 2 or 2A. Patients also had to agree to least one of the following two options: medication switch and medication augmentation. Patients thus only underwent strict randomization among (a) the medication switch options including nortriptyline and mirtazapine, as well as (b) the medication augmentation options including lithium augmentation and triiodothyronine augmentation. Level 4: Patients progressed to Level 4, if they failed to achieve remission in Level 3. Patients were randomized to receive either venlafaxine with mirtazapine augmentation (i.e., “California Rocket Fuel”), or tranylcypromine.
2. COmbining Medications to Enhance Depression outcomes (CO-MED, ClinicalTrials.gov, NCT00590863, [14]) was a single-blind, randomized and multi-center clinical trial that compared the efficacy of escitalopram monotherapy, escitalopram with bupropion augmentation and venlafaxine with mirtazapine augmentation.

We downloaded both datasets from the National Institute of Mental Health Data Archive with a limited access data use certificate awarded to Eric V. Strobl.

### Learning Optimal Outcomes

We used the Supervised Varimax (SV) algorithm to learn outcome measures that maximally differentiate the effects of antidepressants [11]. The SV algorithm considers a supervised principal components or factor analysis model, where treatments ***T*** affect a set of a few factors ***F*** that in turn affect a set of original outcome measures ***Y*** (Figure 1 (a)). We set ***Y*** to the 16 individual items of the Quick Inventory of Depressive Symptomatology - Self Report (QIDS-SR). SV then applies the Varimax rotation to the effect sizes from ***T*** to ***F*** so that each factor is affected by only a small and unique set of treatments. For example, factor ***F***_1_ is affected by all three treatments in Figure 1 (a), but its rotated counterpart 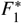 is only affected by *T*_1_ in Figure 1 (b). As a result, the rotated factors ***F***\*, which we also call *optimal outcomes*, now differentiate between treatments.

**Figure 1.**
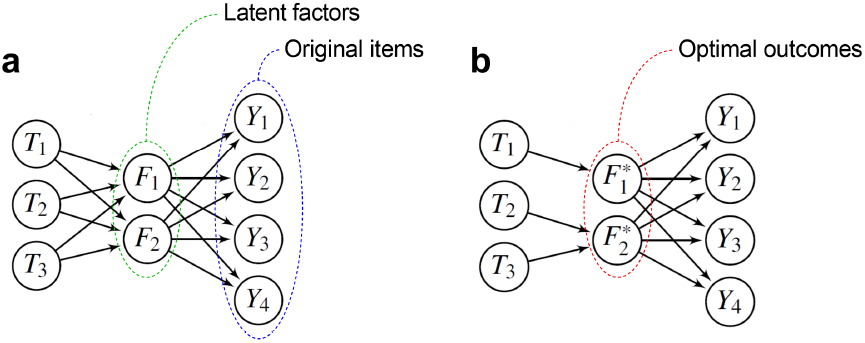
(a) Supervised Varimax (SV) considers a model where treatments treatments ***T*** affect a few factors ***F*** that in turn affect many rating scale items ***Y***. (b) SV learns a set of optimal outcomes ***F***\* such that the effect sizes from ***T*** to ***F***\* are sparse and unique.

### Dependent, Independent and Nuisance Variables

We set all 16 individual items of the Quick Inventory of Depressive Symptomatology - Self Report (QIDS-SR) as the dependent variable for the SV algorithm. We used week 14 data for STAR*D and week 12 data for CO-MED, since CO-MED did not have week 14 data. The independent variables only included binary treatment assignment. We were interested in the differential effects of antidepressants regardless of age, sex and baseline total QIDS-SR score. We therefore partialed out these three variables from each item of the QIDS-SR using ordinary least squares regression before running the SV algorithm.

### Hypothesis Testing

We used permutation tests to control the Type I error rate even after learning the outcomes with SV. We refer the reader to [11] for a detailed description of the permutation tests. We utilized one omnibus test and two post-hoc tests. We always performed 10,000 permutations for each test. The null hypothesis of all three tests corresponds to treatment exchangeability and therefore no differential treatment effect. The alternative hypothesis for the omnibus test refers to at least one differential effect of any treatment in any optimal outcome. The omnibus test uses the *absolute sum* statistic, corresponding to the sum of the absolute effect sizes from ***T*** to ***F***\*.

If we reject the omnibus null hypothesis, then we subsequently perform a post-hoc test for each of the *q* optimal outcomes. The alternative hypothesis of this post-hoc test corresponds to a differential treatment effect in a specific optimal outcome 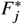 for any treatment. The post-hoc test therefore also uses an *absolute sum* statistic, but corresponding to the sum of the absolute effect sizes from ***T*** to a specific optimal outcome 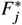. We thus obtain a vector of *q* p-values which we correct using the permutation-based method described in [15] to control the false discovery rate (FDR) below 0.10. We control the FDR because we posit the existence of at least one optimal outcome and thus seek to maximize statistical power, rather than guard against even a single false positive.

If we reject the null hypothesis for a particular factor, then we perform the second post-hoc test, where the alternative hypothesis corresponds to a differential effect between two treatments *T*_*i*_ and *T*_*k*_ in a specific optimal outcome 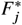. This test uses the *difference* statistic between *T*_*i*_ and *T*_*k*_, corresponding to the effect size of *T*_*i*_ to 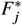 minus the effect size of *T*_*k*_ to 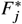. We test all possible treatment pairs within an optimal outcome. Note that the optimal outcomes may contain a few false positives with only FDR control, so we guard against even a single false positive for the treatment pairs by controlling the family-wise error rate (FWER) using the single-step maxT method similar to Tukey’s range test [16].

### Number of Factors

The SV algorithm requires the user to specify the number of factors *q* in ***F***. The algorithm sets *q* = *m* by default, but learning *q* can increase statistical power. We therefore determined *q* by running SV using 2 to 5 factors and then performing the aforementioned omnibus hypothesis test for each number of factors. We set *q* to the number of factors associated with the smallest omnibus p-value.

### Post-Model Selection Inference

Model selection itself can also inflate the Type I error rate, so we always incorporate model selection into the permutation testing by performing selective inference [17]. Let *q* specifically correspond to the number of factors associated with the smallest p-value from the model selection step described in the previous paragraph. In each permutation of each hypothesis test, we shuffle treatment assignment and then perform model selection iteratively until we select *q* factors. Each permutation therefore only corresponds to a shuffle that leads to the selection of *q* factors. Note that model selection requires *u* permutations for *w* candidate numbers of factors (e.g., 4 candidate numbers for 2 to 5 factors) and, if we also perform *u* permutations per hypothesis test, then the described procedure requires around (*wu*)^2^ permutations in total. We speed up the algorithm substantially in the Supplementary Materials by only requiring around 2*wu* permutations.

## Results

### Bupropion

We first applied SV to Levels 2 and 2A of STAR*D to differentiate bupropion from the other antidepressants. Note that patients underwent true randomization only among the medication augmentation and medication switch options. We thus analyzed the two groups separately.

SV differentiated the two augmentation options, including buspirone augmentation and bupropion augmentation, using three optimal outcomes (*n* = 520, absolute sum = 0.322, *p* = 0.013). However, only one of the three optimal outcomes harbored a differential treatment effect between bupropion augmentation and buspirone augmentation (difference = −0.322, *p*_*FWER*_ = 0.005). Visualization of the averaged optimal outcome revealed an increased separation of the treatment responses over time as compared to the total QIDS-SR score (Figure 2 Level 2 top vs. bottom). The optimal outcome corresponded to major depression with increased appetite and weight (Figure 3 (a) red). Bupropion augmentation had a greater therapeutic effect on this optimal outcome than buspirone augmentation.

**Figure 2.**
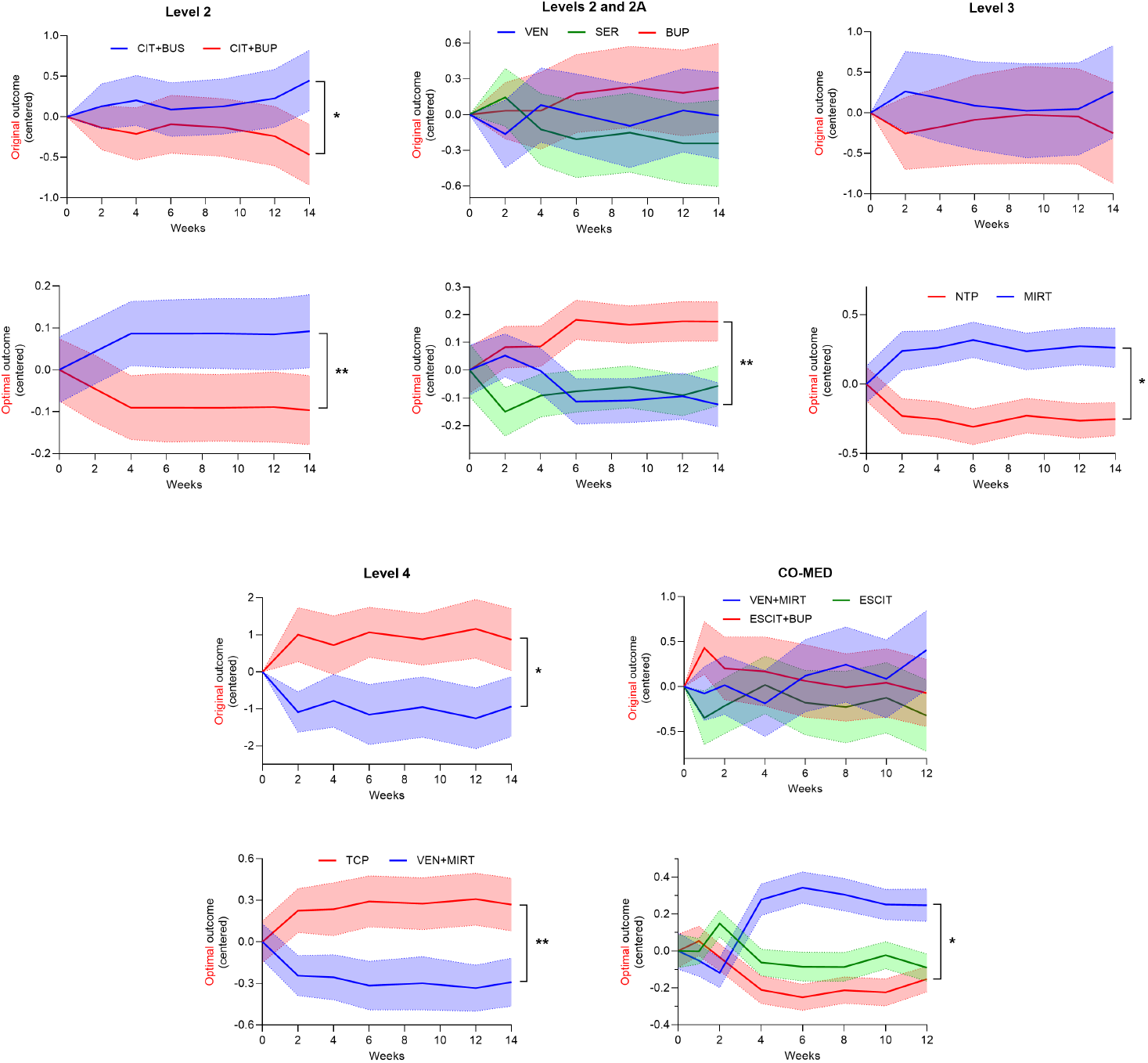
Comparison of the original and optimal outcomes across all time points. Each relevant STAR*D level and the CO-MED trial has a top and a bottom graph; the top graph corresponds to the total score of the original outcomes (i.e., the total QIDS-SR score), and the bottom graph corresponds to the optimal outcome found by SV. We centered the outcomes at each time point, so they have mean zero, in order to focus on treatment differences. If an outcome differentiates between treatments, then the average outcomes of the treatments should separate from each other over time. Notice that the optimal outcomes always differentiated between treatments better than the QIDS-SR total score. Moreover, optimal outcomes always achieved higher statistical power despite model selection and learning of the optimal outcomes. Error bands denote 95% confidence intervals of the mean. CIT denotes citalopram, BUS buspirone, BUP bupropion sustained release, VEN venlafaxine extended release, SER sertraline, MIRT mirtazapine, ESCIT escitalopram. * *p*_*FWER*_ < 0.05, ** *p*_*FWER*_ < 0.01.

**Figure 3.**
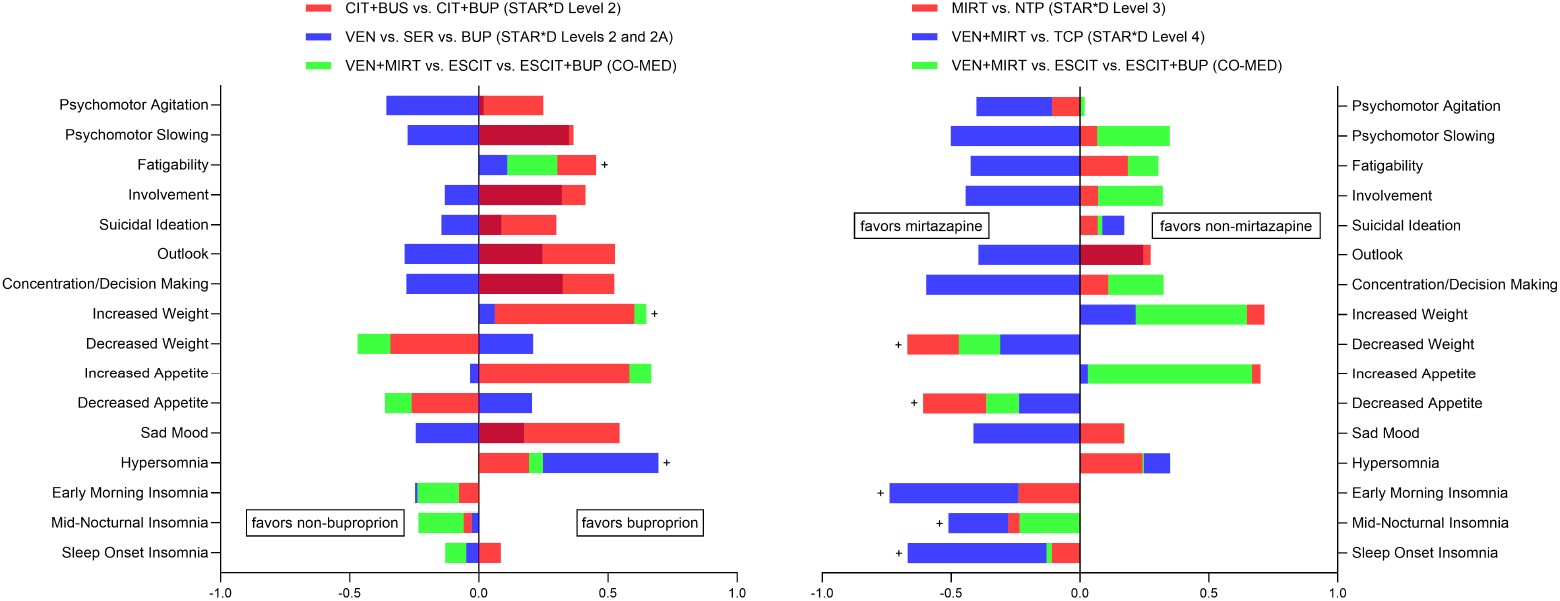
Superimposed bar graphs, where bar heights correspond to the effect sizes of optimal outcomes harboring at least one significant differential treatment effect (*p*_*FWER*_ < 0.05) to individual items of the QIDS-SR. (a) Bar heights to the right correspond to symptoms that respond better to bupropion than the comparator treatment(s). Plus signs highlight symptoms that responded better to bupropion than all other tested treatments. (b) Bar heights to the left correspond to symptoms that respond better to mirtazapine than the comparator treatment(s). Plus signs highlight symptoms that always responded best to mirtazapine.

SV also identified differential treatment effects with bupropion, sertraline and venlafaxine monotherapies from combined Level 2 and 2A data (*n* = 686, absolute sum = 0.666, *p* = 0.040). SV identified five optimal outcomes and detected a differential treatment effect between bupropion and venlafaxine in one of the five outcomes (difference = 0.384, *p*_*FWER*_ = 0.007). The optimal outcome indeed created a much larger separation between bupropion and venlafaxine as compared to the original outcome (Figure 2 Levels 2 and 2A). Bupropion had a larger therapeutic effect on hypersomnia than venlafaxine (Figure 3 (a) blue, rightward bar at hypersomnia).

We replicated the STAR*D results in an independent clinical trial called CO-MED, where patients were randomized to escitalopram with placebo, escitalopram with bupropion and venlafaxine with mirtazapine. SV used three optimal outcomes to learn one optimal outcome that differentiated between the treatments (*n* = 640, absolute sum = 0.396, *p* = 0.047). The optimal outcome differentiated escitalopram with bupropion from venlafaxine with mirtazapine (difference = −0.302, *p*_*FWER*_ = 0.022). Visualization of the optimal outcome revealed a clear separation between escitalopram with bupropion and venlafaxine with mirtazapine over time, a pattern that was not observed in the original outcome (Figure 2 CO-MED). The optimal outcome corresponded to major depression with hypersomnia, increased weight, increased appetite and fatigue (Figure 3 (a) green). Bupropion had a greater therapeutic effect on this optimal outcome than venlafaxine with mirtazapine.

### Mirtazapine

We ran SV on Level 3 of STAR*D to differentiate mirtazapine and nortriptyline monotherapies. The algorithm used two optimal outcomes to identify a differential treatment effect in one of the two optimal outcomes (*n* = 214, absolute sum = 0.401, *p* = 0.048; difference = 0.401, *p*_*FWER*_ = 0.022). The optimal outcome separated mirtazapine from nortriptyline, whereas the original outcome did not (Figure 2 Level 3). Mirtazapine was more effective than nortriptyline in treating decreased weight, decreased appetite and all forms of insomnia (Figure 3 (b) red).

We next compared venlafaxine with mirtazapine against tranylcypromine monotherapy in Level 4 of STAR*D. SV utilized three optimal outcomes (*n* = 102, absolute sum = 0.722, *p* = 0.012) and identified a differential treatment effect between the two treatments in one of the three optimal outcomes (difference = −0.722, *p*_*FWER*_ = 0.004). The optimal outcome separated mirtazapine from tranylcypromine even better than the original outcome (Figure 2 Level 4). The optimal outcome corresponded to major depression with insomnia and decreased appetite. Mirtazapine had a larger therapeutic effect on this outcome than tranylcypromine (Figure 3 (b) blue).

We replicated the results using the CO-MED dataset. As mentioned previously, SV learned an optimal outcome that differentiated venlafaxine with mirtazapine from escitalopram with bupropion (Figure 2 CO-MED). Venlafaxine with mirtazapine was more effective than escitalopram with bupropion and escitalopram with placebo in treatment insomnia, decreased weight and decreased appetite (Figure 3 (b) green).

### Other Results

We completed the analysis of all strictly randomized components of STAR*D by running SV on Level 3 to differentiate lithium augmentation from triiodothyronine augmentation. The algorithm did not detect differential treatment effects with any number of optimal outcomes at even an uncorrected level (*n* = 134, absulute sum = 0.373, *p* > 0.05).

## Discussion

We differentiated mirtazapine and bupropion from each other and from multiple other antidepressants in a comprehensive analysis of two large-scale clinical trials of major depression using the SV algorithm. We replicated the differential effects in all relevant levels of STAR*D, and in an independent trial called CO-MED. Differences between treatment pairs held even after accounting for the learning of the optimal outcomes, model selection and multiple comparisons. We did not detect differential effects between any of the other antidepressants. We thus identified *consistent, significant and specific* differential effects of mirtazapine and bupropion in major depression directly from single clinical trials for the first time.

The recovered differential effects of bupropion and mirtazapine match the unique pharmacodynamics of the two medications. Bupropion is an activating norepinephrine and dopamine re-uptake inhibitor particularly effective for major depression with hypersomnia, fatigue and weight gain [18]. The medication has off-label uses in weight loss and attention-deficit hyperactivity disorder. On the other hand, mirtazapine partially acts as an antihistamine that promotes sleep, reduces nausea and increases appetite at lower doses [19]. Our statistical results therefore match the clinical effects of the medications observed in practice.

Despite the above differences, single clinical trials in psychiatry frequently fail to differentiate mirtazapine and bupropion from each other and from other antidepressants. For example, none of the original STAR*D analyses identified differential treatment effects across multiple levels [1]. The CO-MED study could not differentiate between bupropion augmentation and mirtazapine augmentation [14]. The VAST-D study differentiated bupropion monotherapy from aripiprazole augmentation but failed to differentiate bupropion augmentation and aripiprazole augmentation even with 1522 patients [20]. In contrast, we differentiated bupropion monotherapy and bupropion augmentation from buspirone augmentation, venlafaxine monotherapy and mirtazapine augmentation with only 520-686 patients.

The MIR study enrolled 480 patients but failed to differentiate mirtazapine augmentation from augmentation with placebo [21]. Furthermore, another trial with 218 patients could not differentiate sertraline monotherapy from mirtazapine monotherapy in patients with depression and dementia [22]. One meta-analysis successfully differentiated mirtazapine monotherapy from venlafaxine monotherapy, but the study could not differentiate mirtazapine from other antidepressants without pooling together entire classes of medications involving thousands of patients [5]. We differentiated mirtazapine monotherapy and mirtazapine augmentation from bupropion augmentation, nortriptyline monotherapy and tranylcypromine monotherapy with 102-214 patients. The SV algorithm thus consistently identified differential treatment effects even with substantially smaller sample sizes than many previous studies.

We emphasize that *differentiating* treatments is typically much harder than *predicting* treatment response. No existing machine learning model has successfully *differentiated* treatments, including those involving mirtazapine and bupropion; existing machine learning models only *predict* treatment response using different treatments, but the predictions are often equivocal for any pair of treatments. For example, [6] found that bupropion augmentation predicted a sleep sub-scale better than chance, but the authors did not differentiate bupropion augmentation from mirtazapine augmentation using the sleep sub-scale in CO-MED. [23] found similar treatment responses between bupropion augmentation and mirtazapine augmentation in all identified patient clusters using a deep learning model trained with multiple RCT datasets including STAR*D and CO-MED. [8] successfully predicted remission status better than chance with bupropion but not for any of the augmentation strategies, including bupropion augmentation, in Level 2 of STAR*D. [24] discovered that energy and insomnia are strong predictors of treatment response to bupropion augmentation and mirtazapine augmentation in STAR*D and CO-MED, but the authors could not differentiate the effects of the two treatment strategies. In stark contrast, we differentiated mirtazapine and bupropion without any predictor variables other than treatment assignment with replication in all relevant levels of STAR*D and in CO-MED. We achieved these results because no antidepressant is uniformly better than all other antidepressants for all primary symptoms of major depression. As a result, the total severity score and remission status are coarse outcome measures that significantly lower the statistical power needed to differentiate between treatments. We instead analyzed the *individual items* of clinical rating scales, which increased statistical power significantly despite the learning of the optimal sub-scales, the selection of the number of factors and the corrections for multiple comparisons.

In summary, we consistently identified differential effects of mirtazapine and bupropion by learning optimal outcomes that maximize statistical power. Our approach mimics the strategy adopted by most psychiatrists, where we differentiate treatments based on their unique effects on individual symptoms, rather than based on their unique effects on amalgamated severity scores in patient subgroups.

## Data Availability

All data used in this study is available in the National Institute of Mental Health Data Archive (https://nda.nih.gov/).

## Data Availability

De-identified, individual-level patient data from the STAR*D and CO-MED trials are available at the National Institute of Mental Health Data Archive after approval with a limited access data use certificate.

## Conflicts of Interests

No conflicts exist.

## Acknowledgments

None

## Supplementary Materials

We summarize the permutation procedure in Algorithm 1. We first run SV with model selection using the omnibus hypothesis test in Lines 1 to 6. We save the permutation statistics 𝒮_*q*_ in Line 3 for downstream re-use. We ultimately select the number of factors *q* associated with the smallest p-value in Line 5. Then, for each permutation, we permute treatment assignment and run SV with model selection until we identify an optimal number of factors *q*\* = *q* in Lines 8 to 17. We thus only estimate p-values and control for multiple comparisons in Lines 18 to 22 using the permuted samples where *q*\* = *q*.

Let *w* = |***Q***|. Notice that Lines 1 to 6 only require *wu* permutations. Moreover, we re-use the the permutation statistics S_*q*_ for each *q* ∈ ***Q*** in Line 14. As a result, we only perform *u* new permutations in Line 8. If we only need to repeat Line 11 *O* _*p*_ (*w*) times, then Algorithm 1 requires *wu* + *uO* _*p*_ (*w*) = *O* _*p*_ (*wu*) permutations in total.

### Algorithm 1

Permutation Testing with Selective Inference

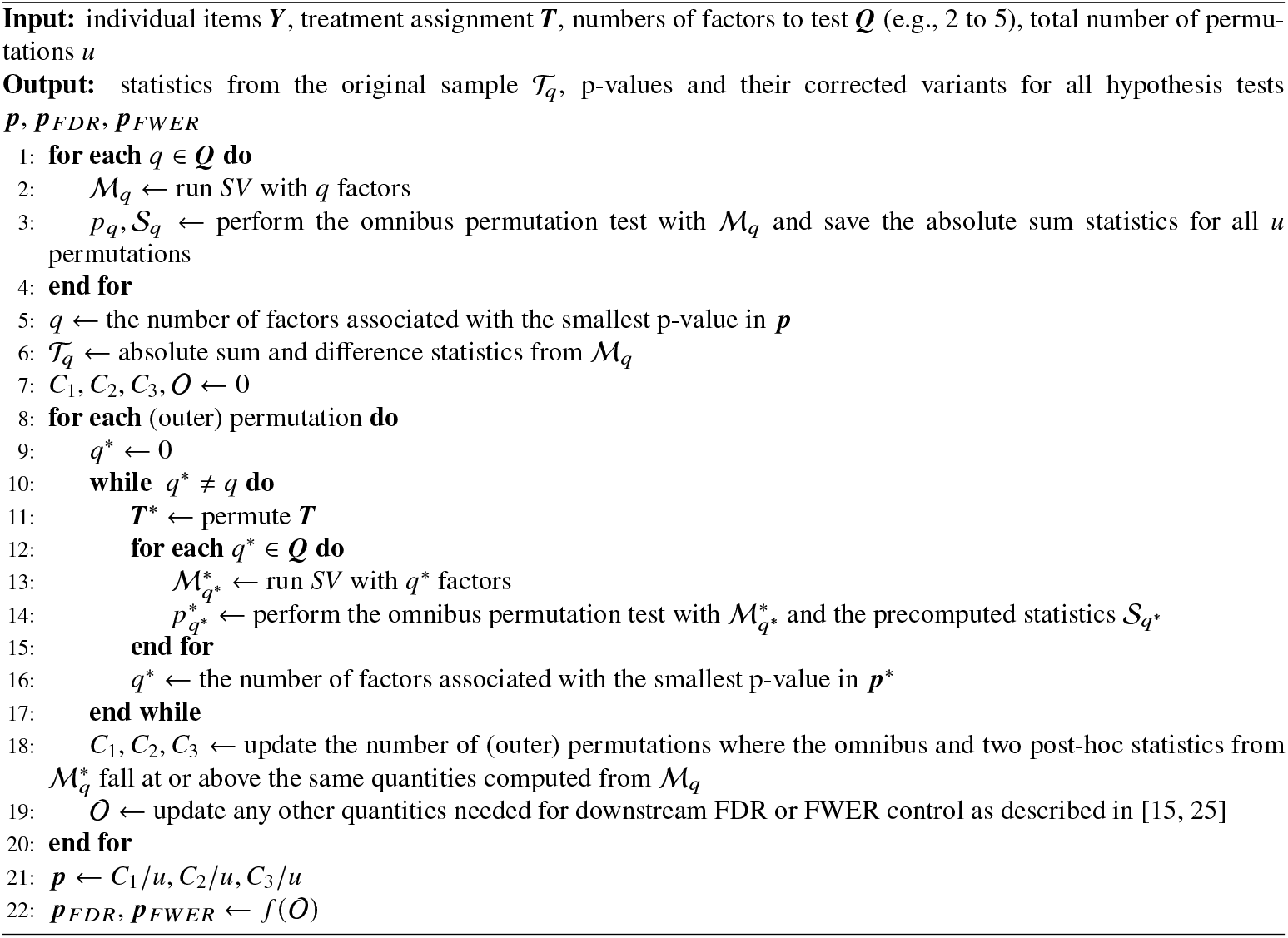

## Notes

### Competing Interest Statement

The authors have declared no competing interest.

### Clinical Trial

NCT00021528, NCT00590863

### Funding Statement

This study did not receive any funding.

### Author Declarations

We downloaded openly available human data from the National Institute of Mental Health Data Archive (https://nda.nih.gov/) with a limited access data use certificate.

### Summary of Updates

Corrected spelling and grammatical errors

